# Clarivate listed nursing journals in 2020: what they publish and how they measure use of social media

**DOI:** 10.1101/2021.04.19.21255561

**Authors:** Roger Watson, Ahtisham Younas, Salma Abdul Rehman, Parveen Azam Ali

## Abstract

**Objectives:** To investigate what the most common types of articles that nursing journals purport to publish are and what they actually publish? And to investigate the extent to which academic nursing journals listed by Clarivate track alternative metrics?

**Methods:** Journals included in the nursing Journal Citation Report journal category in 2019 described as nursing were identified and considered suitable for inclusion in the analysis. Instructions for authors were reviewed online and mention of each type of article identified. The tables of contents of each issue of each journal published during 2019 was examined and the types of articles published were extracted to a spreadsheet into permitted article types and published articles. Likewise, the use of alternative metrics by each journal was extracted to a spreadsheet. Pearson’s and Spearman’s correlation analysis was applied to investigate the relationship between articles permitted and articles published.

**Results:** In the 2020 Journal Citation Report, 123 journals were listed. The most common article type permitted was original research (n=117), followed by review papers (n=116) and discussion papers (n=63). Original research (n=7045); review papers (n=1268); discussion papers (n=1225); editorials (n=793) and commentaries (n=776) were the most commonly published categories of article. Of journals examined, 108 (96.8%) tracked mentions on social media and the Altmetric score was the most commonly use (75%). There was a strong correlation (r=0.73; p=0,002) between the numbers of article permitted and published and a strong correlation (rho=0.86; p<0.001) in terms of the rankings of the permitted and published articles.

**Conclusions:** There is a relationship between the most frequently permitted article types and those published, especially for the most frequent categories of both. Original articles, review papers and discussion papers are the backbone of academic publishing in nursing with original articles vastly outweighing review and discussion papers. Most Clarivate listed journals now use some method of tracking alternative metrics indicating how seriously publishers take their social media profiles.

## INTRODUCTION

The number of academic nursing journals included in the journal impact factor (JIF) league table created and curated by Clarivate (formerly part of Thomson Reuters) shows a net increase annually. Journals are added and removed, and since 2011 it has grown by six journals. The list is always controversial, and the annual release is eagerly awaited by editors and publishers. We do not intend to debate the controversies around JIF in this article; suffice to say that the formula used to calculate the JIF, first used by Eugene Garfield in 1956^1^, is arbitrary and has no meaning outside of its own context. It is, however, an attempt to relate the number of citations a journal receives relative to its size over a defined period. Many alternatives to the JIF have been developed but none are given as much attention and most are, in any case, highly correlated to the JIF^2^. However, whatever controversies surround the use and misuse of the JIF, what remains is the importance for journals of being included or indexed on the list. Along with other listing such as PubMed, Clarivate listing in itself is a mark of prestige and, to be listed, Clarivate examine a range of factors including how international the journal is in terms of authorship and editorship; the extent to which is follows international academic publication standards and follows publication ethics. Journals must not excessively self-cite their own material or encourage this in any way, or they risk – as has happened^3^ – being removed from the listing.

Our aim in this study was to examine the academic nursing journals listed in 2020 in the Clarivate listing of those currently awarded a JIF. We were interested to investigate, generally, the scope of what these journals purport to publish and to compare it with the scope of their actual output. In addition, while these journals all have a JIF and the Clarivate list contains other citation-based metrics, we were also interested to discover how much use was made of alternative metrics, which measure the extent to which the contents of these journals are referred to in a range of social media sites.

## BACKGROUND

The publishers and editors of academic journals pay close attention to the performance of their journals as do potential authors^4,5^. The reasons why publishers and editors pay attention to their performance is to draw comparisons with their competitors and to consider how to improve that performance. Authors give this attention because, generally, it is considered better by authors to publish is higher performing journals. This is most commonly due to pressure from academic employers – universities and research bodies – to publish more^6^ and be seen to be published in the most prestigious places and, while the extent to which this pressure is applied across the world varies, it is unusual anywhere for some measure of journal performance not to be taken into consideration by authors.

Naturally, the most commonly referred measure of journal performance is the journal JIF, referred to in the Introduction above. The appeal of the JIF lies mainly in its simplicity and the fact that it is widely used. On the other hand, it is widely misunderstood and, while most (but by no means all) journal editors can explain its calculation and what it means; few authors can do this precisely^7^. First, it is not a measure of any kind of impact on research and practice, as widely misunderstood. It is purely a measure of citation activity to one journal and, arbitrarily, it is a measure of the number of citations in one year to the articles published in the previous two years. It come as a surprise to many authors that citation within the year of reporting the JIF does not contribute to the JIF^8^. The failings of the JIF are well known, and these include the fact that only a small percentage of the article published in a journal contribute towards the JIF, there is always a ‘tail’ of articles in any journal that never get cited. Thus, the JIF tells us nothing about the performance of most articles in a journal. The debate about the uses and abuses of the JIF have been rehearsed elsewhere and a consideration of the JIF is not the focus of this article.

Citation based alternatives to the JIF have almost universally failed to make inroads into the debate about journal performance^9^. One attractive feature of the JIF is the relative ease with which it can be calculated: the number of citations in the reporting year to the articles published in the previous two years divided by the number of articles published in the previous two years. All the alternatives such as Eigenfactor score of the Scimago ranking attempt to account not only for the number of citations but where those citations take place. Thus, they provide weightings for more prestigious journals but problem with the formulae for calculating these alternative indices is that they are inordinately complicated. Another feature which tends to render them redundant is that these measures, being citation based, all correlate highly with the JIF and therefore do not provide much additional information about the relative performance of journals^2^.

While the primacy of the JIF remains, although frequently discussed and questioned, and alternative citation-based metrics continue to be considered, a phenomenon took place which has led to the possibility for an alternative type of metric; one which has the potential to be more meaningful and which is growing in importance. The phenomenon is social media, and the new measures can be collectively described as alternative metrics. There is already interest in how journals are mentioned in social media^10^ and advice on how best to make use of this^11^. There is a range of alternative metrics with two being much more common than the others. The original alternative metric was the Altmetric© and this was used by a range of publishers and was the main measure until the publisher Elsevier introduced the PlumX© metric. Both the Atmetric score and the PlumX score are run as commercial concerns and they are, demonstrably, very similar^12^. Both systems use mentions in a range of social media sites such as online newspapers, Twitter© and blogs to generate their scores. Not all social media sites are considered to be equal and, for example, under the Altmetric system a mention in an online newspaper generates a higher score than a mention on Twitter. The PlumX metric includes mention in Mendeley, which is not used in the Altmetric score and it is notable that Elsevier – the owners of PlumX – also own Mendeley. These alternative metrics are updated in real time, which is a very useful advantage over citation-based metrics which are update annually. They are article specific and an alternative metric score is not awarded to a journal. A particular feature of both the Altmetric and the PlumX systems is that they both use attractive and recognisable logos which appear on the landing page for an article with a number attached indicating the score. Using both logos it is possible to obtain more details than the score. With the Altmetric logo the reader can hover over the logo with the mouse cursor and to see a popup with a basic indication of the social media sites where the article has been mentioned. Clicking on ‘Further information’ provides a more detailed breakdown including a world map showing where the mentions have taken place and the type of citation, for example, ‘Members of the public’ or ‘Scientists’. Next to the PlumX logo clicking on ‘Further information’ provides a breakdown of where mentions have taken place. The inclusion of Mendeley allows the reader to see in which other articles the article has been cited.

A cursory inspection of the Clarivate list of academic nursing journal shows that other metrics are used by a few journals, specifically Metrics and Dimensions©. We were unable to obtain any independent information about Metrics but, where it is used, Metrics provides a table of where the article has been mentioned and to our best knowledge, these include: Crossref; Google Scholar; and Scopus. Dimensions is metrics based but incudes reference in patents and has an attractive logo which serve a similar function to the Altmetric logo.

Journal metrics of whatever kind are related to what the journals publish, the journal content. The content of journals varies, and all authors know this as they target specific journals for particular kinds of manuscript+98. Moreover, the content of journals evolves as publishers and editors review their contents, usually for performance in terms of metrics, hits on their webpages and downloads. For example, some academic nursing journals will publish concept analyses, while others eschew them. It is also known that some kinds of articles attract more citations than others. For example, it is widely known that review articles tend to me more highly cited than other kinds of articles^13^. This is one of the only identifiable patterns in academic publishing and, while other articles can be highly cited, usually based on their own merits, there is usually no discernible pattern. Therefore, it should be of interest to those who use academic nursing journals to compare and contrast the contents of these journals. It should also be of interest to catalogue what the most common types of articles that journals purport to publish are and what they actually publish?

### Research Questions

To investigate the above phenomena, the research questions guiding this study were:

1. What do academic nursing journals listed by Clarivate purport to publish?
2. What do academic nursing journals listed by Clarivate actually Publish?
3. To what extend do academic nursing journals listed by Clarivate track alternative metrics?

## METHODS

We used Clarivate Analytics’ 2020 Journal Citation Report (JCR) based on data for 2019 to access the information about nursing journals indexed in Web of Science with a JIF. Specifically, all journals included in the JCR journal category described as nursing were identified and considered suitable for inclusion in the analysis. The JCR includes various indicators of journal performance (such as JIF) and ranks journals in two broad categories of type of science: the Science Citation Index Expanded (SCIE) and the Social Science Citation Index (SSCI). There were 123 journals in the 2019 JCR nursing category.

To identify what journal say they publish, the instructions for authors were reviewed online and mention of each type of article identified. The instructions were then downloaded and each of these documents was carefully read and the types of articles permitted were extracted to a spreadsheet. To explore what journal published, the tables of contents of each issue of each journal published during 2019 was examined and the types of articles published were extracted to a spreadsheet. Likewise, the use of alternative metrics by each journal was extracted to a spreadsheet. The data were then enumerated and recorded under each type of article and each type of alternative metric and tabulated. Any anomalies were checked by two authors.

In terms of article types, for ease of presentation and interpretation, in recording and reporting what journals permit and publish some categories of papers were collapsed. For example, meta-analyses and other systematic reviews are all reported as ‘Systematic reviews’ and the category ‘Original research’ included qualitative, quantitative and mixed-methods studies and methodological papers, which 14 journals permitted. are included under discussion papers. Data were entered into SPSS version 27.0 for analysis using Pearson’s and Spearman’s correlation.

## RESULTS

In the 2020 Journal Citation Report, 123 journals were listed. Table 1 shows, in order of frequency what journals purport to publish. We report 15 types of articles that journals said they permitted as submissions. The most common article type permitted was original research (n=117), followed by review papers (n=116) and discussion papers (n=63).

**Table 1.** Categories of articles permitted in order of frequency.

| **Category of article** | **N** |
| --- | --- |
| Original research | 119 |
| Review papers | 116 |
| Discussion papers | 92 |
| Correspondence | 51 |
| Editorials | 36 |
| Case studies | 33 |
| Commentaries | 32 |
| Quality improvement | 27 |
| Short reports | 17 |
| Policy analyses | 11 |
| Protocols | 6 |
| Pilot studies | 4 |
| Concept analysis | 4 |
| Book/media reviews | 3 |
| Expert interview | 1 |

We the report these 15 categories of manuscript published in journals as shown in Table 2. The top three categories mirrored the permitted types of manuscripts as follows: original research (n=7045); review papers (n=1268) and discussion papers (n=1225). Editorials (n=793) and commentaries (n=776) were the next most commonly published categories of article. Editorials were ranked fifth in the types of permitted manuscript (Table 1; n=36) but were ranked fourth in the type of article published (n=793) followed by commentaries (n=776). It should be noted that, while two journals reported that they permitted poetry, we found no examples in the period examined. Figure 1 represents the relative percentages of the types of articles in a funnel plot; we used this type of plot due to the fact that the total number of article types exceeded 100%.

**Table 2.** Categories of articles published in order of frequency.

| **Type of article** | **N** |
| --- | --- |
| Original research | 7045 |
| Review papers | 1268 |
| Discussion papers | 1225 |
| Editorials | 793 |
| Commentaries | 776 |
| Pilot studies | 173 |
| Correspondence | 142 |
| Short reports | 125 |
| Quality improvement | 87 |
| Concept analysis | 70 |
| Case studies | 59 |
| Protocols | 58 |
| Policy analyses | 48 |
| Book/media reviews | 34 |
| Expert interview | 21 |
| **Total** | **11924** |

**Figure 1:**
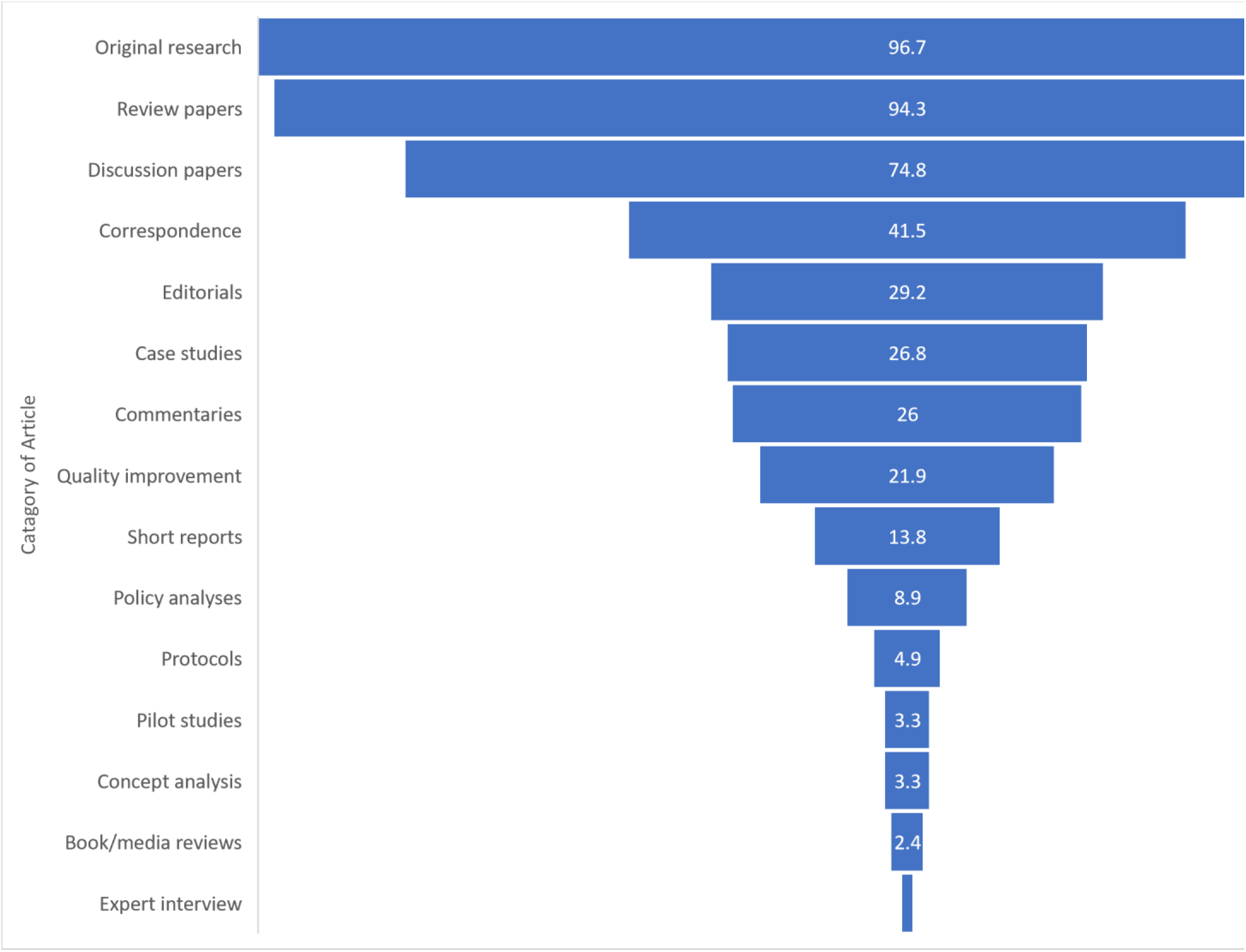
Categories of articles permitted in order of frequency (%)* *NB: the total % is > 100 as journals publish > type of article

Table 3 shows the frequency with which journals reported the use of alternative metrics. Of the 123 journals examined, 108 (96.8%) tracked mentions on social media. By far the most common method of tracking social media use was the Altmetric (n=75) followed by the Elsevier journals’ use of PlumX (n=29). Only two journals use each of Metrics and Dimension. Figure 2 represents the relative percentages (total = 100%) in a pie chart.

**Table 3.**
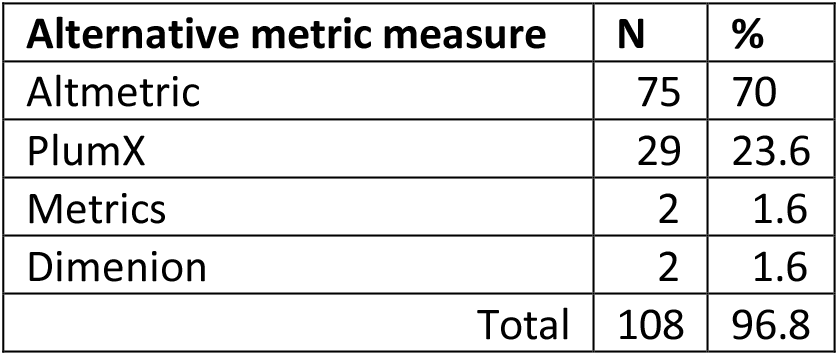
Alternative metric measures used by journals.

| **Alternative metric measure** | **N** | **%** |
| --- | --- | --- |
| Altmetric | 75 | 70 |
| PlumX | 29 | 23.6 |
| Metrics | 2 | 1.6 |
| Dimenion | 2 | 1.6 |
| Total | 108 | 96.8 |

**Figure 2.**
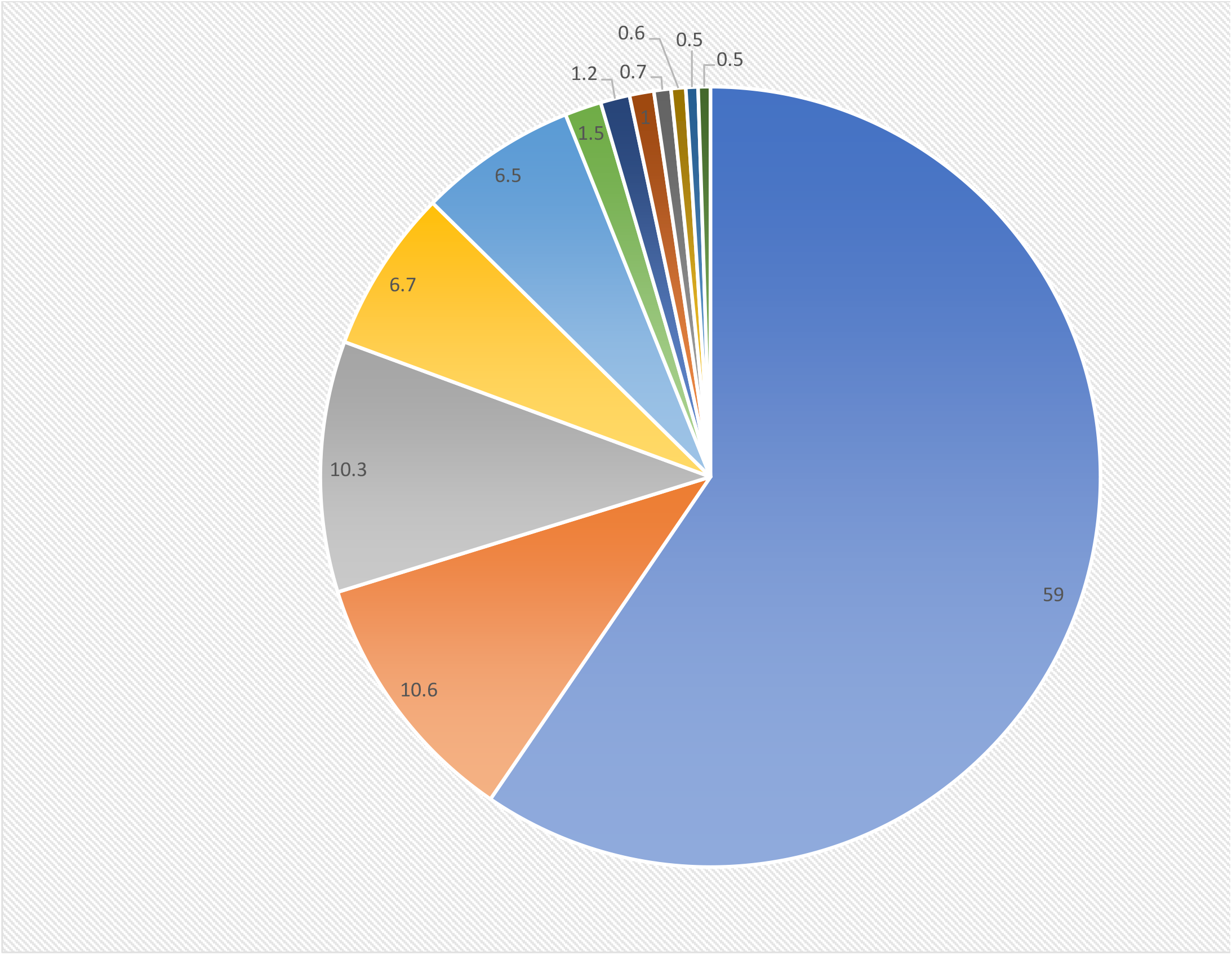
Categories of articles published in order of frequency (%)

Pearson’s correlation (r=0.73; p=0,002) between the number of articles permitted and published and Spearman’s correlation (rho=0.86; p<0.001) in terms of the rankings of the permitted and published articles were both strong.

## DISCUSSION

We set out to investigate what academic nursing journals say they publish and what they actually publish. We also examined the use of methods of tracking mentions of journal content on social media. The most obvious conclusion from our results is that not all journals publish the same range of articles. Also, there is an obvious relationship, between the most frequently permitted article types and those published, especially for the most frequent categories of both. There is remarkable congruence between the top three categories of article permitted and published: original articles; review papers and discussion papers, respectively. These three types of article are, clearly, the backbone of academic publishing in nursing with original articles vastly outweighing review and discussion papers. This is hardly surprising due to the amount of original research taking place as an outcome of funded research and masters and doctoral projects being caried out in hospitals and universities. Reviews and are dependent on original research and seek, periodically, to synthesise original research with a view to providing best evidence, identify gaps and formulate novel research questions^14^. To some extent, discussion papers are also – but not completely – dependent on original research; some seek to take current issues or theoretical and methodological developments and set an agenda for further research and discussion.

Editorials are the next most commonly published type of article and the importance of editorials to nursing journals is something that has been considered^15^. Editorials serve a range of purposes such as allowing the editors of a journal to promote the content of the journal of to promote their views. But they are also used to indicate when changes have taken place in journal content^16^, to discuss issues related to journal content^17^ and to request particular types of content^18^. Also, editorials, while not included in the denominator for the calculation of the JIF, citations to editorials within the two-year window for calculating the JIF are added to the numerator and these constitute ‘free cites’. As such, these can be very valuable to editors and publishers as a way of increasing their JIFs. In this light, it can be very valuable to permit critical discussion of published articles, whether this is positive or negative^19^. In a similar light, correspondence and commentaries can also contribute to free cites in the journal, although these will frequently – but not always – be published alongside or within the same year as publication as the articles to which they refer and, thereby, may not contribute to the calculation of the JIF. Correspondence was the fourth most common category of permitted article but was only seventh, behind editorials and commentaries and pilot studies in terms to frequency of publication.

Concept analysis articles were only permitted by four journals. There has been a move away from publishing concept analysis articles by at least one journal in the list – the *Journal of Advanced Nursing* – and this led to some controversy^20,21^. It appears that, despite the low number of journals permitting concept analysis articles that there is still a desire to publish them as 70 articles were published in the period of the study. This suggest that these articles are clustered in a very small number of journals. The same could be said for protocol articles, which were only permitted by six journals but of which 58 articles were published. Publishing protocols has only relatively recently been encouraged by journals^22^ and has arisen from the increasing requirement to register clinical trials and to specify study protocols as required of academic journals with publishers who are signatories of the AllTrials Campaign^23^.

Increasingly, academic nursing journals have ceased to publish book reviews, which were once very comment^24^ and this is reflected in the fact that only three journals permit them and, concomitantly, publish them. It is also apparent then expert interviews play a very small part in publishing in academic nursing journals. The above review of content is not comprehensive but highlights some main points of interest.

It is hardly surprising that both the raw and the ranked correlation between articles permitted and published were strong. However, for future similar studies in nursing and in other subjects a baseline has been established whereby trends may be measured and compared.

### Alternative metrics

Most Clarivate listed journals now use some method of tracking alternative metrics which is an indication of how seriously publishers take their social media profiles as represented by reference on a range of social media sites to the content of their journals. While there is some evidence for a relationship between social media mentions and citations^5,25,26^, the evidence is not strong and compounded by the fact that highly cited articles may, subsequently, be the ones that receive greater attention on social media. Thus, is it hard to discern cause from effect. However, the use of alternative metrics is not solely concerned with its relationship to and potential for increasing citations and the impact factor; alternative metrics offer a wider insight into the impact of the work published in a journal and, especially, beyond the scientific community, although this aspect of the use of alternative metrics, with one exception^5^, has not been widely or systematically studied.

### Limitations and recommendations

Our study was restricted to nursing journals and it would be useful to compare the situation investigated here with other cognate subjects such as medicine and allied health. Our study was purely descriptive and did not formally investigate relationships between any of the variables; as such it aimed only to provide baseline information for further investigations. We already know what factors lead to greater citations in academic journal, for example publishing review article and also methodological articles^27^. We strongly recommend that future studies investigate how the range of what is published in academic journals relates to the use of and success with alternative metrics.

For convenience we combined some categories of articles. As such we made no distinction between, for example, qualitative and quantitative studies and systematic reviews with and without meta-analysis. Future studies could achieve greater granularity and could also investigate trends in the publication of different types of article and how this relates to different trends in research and the measurement of metrics,

## Data Availability

Data are available on request from authors

